# Epigenome-Wide Association Study of the Upper-Airway Microbiome Involved in Asthma Exacerbations

**DOI:** 10.64898/2026.10.02.26364005

**Authors:** Erick Castillo-Vargas, Mario Martin-Almeida, Anne K. Bozack, Elsa L. Garcia, Elena Martin-Gonzalez, Ruperto González-Pérez, José M. Hernández-Pérez, Olaia Sardón-Prado, Paloma Poza-Guedes, Elena Mederos-Luis, Paula Corcuera-Elosegui, Inmaculada Sánchez-Machín, Leyre López-Fernández, Sheryl L. Rifas-Shiman, Simone Hashimoto, Susanne J. H. Vijverberg, Anne H. Neerincx, Heike Buntrock-Döpke, Mario Gorenjak, Antoaneta A. Toncheva, Susanne Harner, Susanne Brandstetter, Christine Wolff, Uroš Potočnik, Michael Kabesch, Aletta D. Kraneveld, Jesús Villar, Marie-France Hivert, Emily Oken, Diane R. Gold, Mahmoud I. Abdel-Aziz, Anke H. Maitland-van der Zee, Andres Cardenas, Fabian Lorenzo-Diaz, Maria Pino-Yanes, Javier Perez-Garcia

**Author notes:** **Corresponding author:** Javier Perez-Garcia, PhD. Genomics and Health Group, Department of Biochemistry, Microbiology, Cell Biology and Genetics, Universidad de La Laguna (ULL), La Laguna, Tenerife, Spain. Department of Epidemiology and Population Health, Stanford University School of Medicine, Stanford, CA, USA.

## Abstract

**Rationale:** The role of DNA methylation (DNAm) markers in the upper-airway microbiome related to asthma exacerbations despite inhaled corticosteroid (ICS) treatment is unexplored.

**Objectives:** To identify whole-blood DNAm markers associated with the upper-airway microbiome related to asthma exacerbations despite ICS treatment.

**Methods:** We analyzed salivary, nasal, and pharyngeal microbiome in 242 asthma patients (GEMAS) for discovery. We tested associations between whole-blood DNAm at 703,505 CpGs and 21 exacerbation-related microbiome traits. Replication for the nasal microbiome was evaluated in 272 (Project Viva) and 66 (SysPharmPediA) children with and without asthma. We analyzed differentially methylated regions (DMRs), gene enrichment, associations with blood gene expression (RNA-seq, n=119), and DNAm predictors of plasma proteins.

**Measurements and Main Results:** Whole-blood DNAm at 73 CpGs and 550 DMRs was associated with 17 exacerbation-related microbiome traits (false discovery rate [FDR]<0.05), including genes implicated in immune and inflammatory responses and the 17q12-q21 asthma locus *ORMDL3*. Most CpGs were associated with *Campylobacter* (pharyngeal) and *Leptotrichia* (nasal) abundance. Ten DNAm markers were replicated (*p*<0.05), including the corticosteroid insensitivity-related gene *PTPRR* associated with nasal bacterial richness. Enrichment analyses identified genes related to diabetes, obesity, smoking, TGF-β, MAPK, and Wnt signaling and regulated by trichostatin A, NF-κB, and the glucocorticoid receptor (FDR<0.05). The CpG cg18994033, associated with pharyngeal *Tannerella*, was associated with *ITM2C* whole-blood expression. Nine DNAm-predicted plasma proteins involved in inflammation, tissue remodeling, and TGF-β signaling were associated with seven exacerbation-related microbiome traits, including *Streptococcus* and *Selenomonas*.

**Conclusions:** Whole-blood DNAm markers implicated in inflammation, immunity, and asthma comorbidities were associated with the upper-airway microbiome linked to asthma exacerbations.

## INTRODUCTION

Asthma affects over 300 million individuals and ranks as the 34^th^ leading cause of disease burden globally.^1,2^ Asthma patients may experience worsening episodes of the disease, known as exacerbations, which lead to reduced quality of life and major complications such as intubation, hospitalization, or even death.^2^ Although asthma is commonly controlled with inhaled corticosteroids (ICS), 17% of patients have difficult-to-treat asthma, and 3.7% have severe asthma with treatment unresponsiveness.^2^ Given that asthma exacerbations result from the interaction of genetic and environmental factors,^3^ there is increasing interest in studying the role of the microbiome and epigenome.^4,5^

Epigenetic modifications can regulate gene expression without altering the underlying genetic sequence.^4^ DNA methylation (DNAm) is the most studied epigenetic marker, consisting of the methylation of 5’-cytosine-guanine-3’ dinucleotides (CpG). DNAm at thousands of CpGs and differentially methylated regions (DMRs) has been associated with asthma and exacerbations.^4,6,7^ Most asthma-related epigenome-wide association studies (EWAS) have focused on whole blood, which may capture systemic immune and inflammatory mechanisms, and nasal epithelia, which provide information on environmental exposures and airway pathophysiology.^6^

The human microbiome encompasses the community of genetically characterized microorganisms that inhabit the human body.^5^ The oral and nasopharyngeal microbiome have been described as proxies for lung microbiome dysbiosis that may influence asthma pathogenesis.^8^ In a previous study, we identified 18 bacterial genera and three α-diversity indices from the upper airways associated with asthma exacerbations despite ICS treatment.^8^ Through microbiome genome-wide association studies (mbGWAS), we identified host genetic variants associated with these upper-airway microbiome traits, detecting genes involved in inflammation and asthma comorbidities and risk factors (e.g., obesity, reflux esophagitis, and smoking).^9^

Epigenetic modifications are increasingly recognized as key mediators of host-microbiome interactions, with bacteria-produced metabolites shaping the host epigenetic landscape through multiple mechanisms.^6^ However, integrative microbiome epigenome-wide association studies (mbEWAS) remain scarce, with only two conducted to date involving the respiratory microbiome in adolescents and infants with severe bronchiolitis.^10,11^ They identified associations between nasal and blood DNAm and the respiratory microbiota, including genes involved in hedgehog signaling, inflammation, and immune functions, as well as circulating proteins linked to the microbiome-related differences in DNAm.^10,11^ In addition, higher *H. influenzae* abundance has been associated with DNAm age deceleration and increased asthma risk.^10^ Nevertheless, no mbEWAS has focused on asthma patients.

Therefore, we aimed to identify DNAm markers in whole blood associated with upper-airway microbiome traits related to asthma exacerbations despite ICS treatment. We conducted mbEWAS in 242 European asthma patients and assessed replication in 338 ethnically diverse children from two independent cohorts. We examined potential underlying biological pathways, functional consequences on whole-blood gene expression, and DNAm predictors of plasma protein levels.

## METHODS

An extensive Methods section is reported in the **Online Supplement**.

### Study Populations

In the discovery phase, we analyzed 242 individuals with asthma from the Canary Islands (9-82 years) and Basque Country (8-15 years) enrolled in the GEMAS study with available data from the nasal, salivary, and pharyngeal microbiome and blood epigenome. In the replication phase, we analyzed children with nasal microbiome and blood DNAm data, including 272 adolescents with and without asthma from the Project Viva cohort and 66 children with moderate-to-severe asthma from the SysPharmPediA study. All studies were approved by the corresponding ethics committees of the participating centers (GEMAS: approval 29/17 for the hospitals in the Canary Islands and PI2019077 for the University Hospital Donostia; SysPharmPediA: approval 18-1034-101 for centers in Germany, PI2015075[SO] in Spain, NL55788.041.15 in the Netherlands, 0120-569/2017/4 in Slovenia, and Dnr 2010/1336-31/3 and 2019-00546 in Sweden; Project Viva: protocols approved by the Harvard Pilgrim Health Care Institutional Review Board), and participants provided written informed consent or assent, as appropriate.

### Microbiome Profiling

Microbiome data were generated by targeted sequencing of the 16S rRNA gene (V3-V4 region) in salivary, nasal, and pharyngeal samples in GEMAS. Sample collection and quality control (QC) of microbiome data is described elsewhere,^8^ ensuring the removal of non-bacterial sequences, contaminants, and low-quality reads. After QC, ASVs were collapsed at genus level, and data were log-transformed after adding a pseudocount of 1 for subsequent analyses. The nasal microbiome was profiled in SysPharmPediA (V3-V4 region) and Project Viva (V1-V3 region) as described in the **Supplementary Methods**.^12^

### Processing of Epigenomic Data

Whole-blood DNAm in >800,000 CpGs was measured in all cohorts using the Infinium MethylationEPICv1.0 BeadChip (Illumina). A detailed description of sample processing and QC is provided elsewhere and summarized in the **Online Supplement**.^7,13,14^ Briefly, we filtered probes with 1) low-quality data, 2) high missingness rate, 3) multimodal distribution of beta values, 4) cross-reactive, 5) polymorphic, and 6) located on sex chromosomes. We removed samples with 1) low-quality data, 2) high missingness rate, 3) discordance between epigenetic and reported sex, and 4) potentially cross-sample contaminated. DNAm was expressed as M-values and corrected for batch effect. Blood cell type composition was estimated from DNAm data using the ReFACTor algorithm.^15^

### Statistical Analysis

#### Microbiome Epigenome-Wide Association Studies

In the discovery, we tested associations between 703,505 CpG sites and three α-diversity indices and 18 bacterial genera previously related to asthma exacerbations in GEMAS.^8^ Models were adjusted for age, sex, genetic ancestry, cell-type heterogeneity, and batch effect. Analyses were performed separately in Canary Islands and Basque Country subpopulations and meta-analyzed. Significance was declared at a false discovery rate (FDR)<0.05. We conducted sensitivity analyses for pseudocount addition (0.1, 0.5, and 1) prior to log-transformation of the microbiome data and for potential confounders of the upper-airway microbiome previously identified in GEMAS.^8^ For nasal samples, these included body mass index (BMI) categories, season of sampling, and smoking within 30 minutes prior to sample collection (in adults), whereas for pharyngeal samples they included antibiotic prescription in the last two months, cavities, and liquid intake within 30 minutes prior to sample collection. Sensitivity analyses for antibiotics prescription were conducted for all biological sample types.

#### Differentially Methylated Regions

DMRs were identified using ipDMR^16^ and comb-p^17^. Regions were defined by aggregating CpGs within a 1 kb window, and only significant DMRs (FDR<0.05) containing at least 2 CpGs were retained. We focused on DMRs detected using the two software with an overlap >50% of genomic range. Gene annotation was provided using the *rGREAT* package.^18^

#### Enrichment Analyses

We conducted gene-set enrichment analyses (GSEA) for traits, gene ontologies, biological pathways, and drug signatures using Enrichr^19^. We analyzed gene annotations from suggestive CpGs (*p*<5×10^-5^) from the mbEWAS. Significant enrichment terms were declared using an FDR<0.05. To ensure the robustness of associations, we prioritized enrichment terms that remained significant (*p*<0.05) after reassessing GSEA by varying alternative input cutoffs for gene selection (*p*<5×10^-4^ and *p*<5×10^-6^). Additionally, we conducted stratified enrichment analyses for saliva, pharyngeal, and nasal samples.

#### Replication Analyses

CpGs and enrichment terms significantly associated with the nasal microbiome in GEMAS were followed up for replication in Project Viva (n=272) and SysPharmPediA (n=66), following a similar methodology as in the discovery. Details about models and covariates are described in the **Supplementary Methods**. Replication was declared at *p*<0.05, and the same effect direction for CpGs. For DMRs replication, we tested associations between the first principal component (PC_1_) of M-values across all CpGs from each DMR with microbiome traits using linear regression, adjusting for the same covariates as in the discovery mbEWAS.

#### Quantitative Trait Analyses

Whole-blood RNA-seq data were generated for 119 GEMAS participants with DNAm data. We conducted cis-expression quantitative trait methylation (eQTM) analyses testing associations between CpGs identified in mbEWAS and blood expression of nearby genes (±100 kb), while adjusting for age, sex, genetic ancestry, cell composition, asthma, and surrogate variables. Significance was declared at FDR<0.05. We conducted *in silico* associations with genetic variation through methylation quantitative trait loci analysis (meQTL) in whole-blood, using the MeQTL EPIC^20^ database, and with blood gene expression (eQTMs) using HELIX Project^21^ and BIOS QTL.^22^

#### Assessment of CpGs Previously Related to ICS Response or Asthma Exacerbations

We examined whether 22 whole-blood CpGs identified by previous EWAS of ICS response or asthma exacerbations (**Table S2**) were associated with exacerbation-related bacteria. Significance was set at FDR<0.05.

#### DNAm Predictors of Plasma Proteins

We estimated plasma levels of 109 proteins using DNAm predictors. We tested associations between DNAm-predicted proteins and the exacerbation-related microbiome traits using regression models adjusted for the same covariates as in the discovery. Results were meta-analyzed across the Canary Islands and Basque Country subgroups. Significance was defined at FDR<0.05.

## RESULTS

### Study Populations

We analyzed 242 participants with asthma from GEMAS in the discovery phase, including 191 individuals from the Canary Islands and 51 children from the Basque Country. Their characteristics are summarized in **Table 1** and **Table S1**. Briefly, the mean age was 38.4±21.7 years, 61.6% were female, and 31.1% had obesity. All participants were European, and 53.7% experienced asthma exacerbations in the previous 12 months. In the replication phase, we analyzed 66 children with moderate-to-severe asthma predominantly European from the SysPharmPediA consortium (mean age: 11.5±2.9 years, 40.9% female) and 272 adolescents, predominantly White and with 17.3% having asthma, from the Project Viva cohort (mean age: 13.0±0.7, 47.1% female) (**Table 1**).

**Table 1.** Descriptive summary of participants analyzed in this study.

| Variable | Discovery |  |  | Replication |  |  |
| --- | --- | --- | --- | --- | --- | --- |
|  | N | GEMAS | N | Project Viva | N | SysPharmPediA |
| Age (years) | 242 | 38.4 ($\pm 21.7$) | 272 | 13.0 ($\pm 0.7$) | 66 | 11.5 ($\pm 2.9$) |
| Sex (female) | 242 | 149 (61.6) | 272 | 128 (47.1) | 66 | 27 (40.9) |
| Race and Ethnicity | 225 |  | 272 |  | 64 |  |
| European/Non-Hispanic White |  | 225 (100.0) |  | 167 (61.4) |  | 49 (76.6) |
| Latino/Hispanic |  | 0 (0.0) |  | 33 (12.1) |  | 6 (9.1) |
| African/Black |  | 0 (0.0) |  | 47 (17.3) |  | 3 (4.5) |
| Other |  | 0 (0.0) |  | 25 (9.2) |  | 6 (9.4) |
| Asthma | 242 |  | 272 |  | 66 |  |
| No asthma |  | 0 (0.0) |  | 190 (69.9) |  | 0 (0.0) |
| Ever asthma |  | 0 (0.0) |  | 35 (12.9) |  | 0 (0.0) |
| Current asthma |  | 242 (100.0) |  | 47 (17.3) |  | 66 (100.0) |
| Asthma exacerbations* | 242 | 130 (53.7) | 70 | 11 (15.7) | 66 | 38 (57.6) |
| BMI categories | 228 |  | 270 |  | 65 |  |
| Underweight/Normal Weight |  | 77 (33.8) |  | 175 (64.8) |  | 42 (64.6) |
| Overweight |  | 80 (35.1) |  | 73 (27.0) |  | 13 (20.0) |
| Obesity |  | 71 (31.1) |  | 22 (8.1) |  | 10 (15.4) |
| FEV<sub>1</sub> (z-score) | 228 | -0.5 ($\pm 1.2$) | 259 | 0.0 ($\pm 1.1$) | 65 | 0.0 ($\pm 1.3$) |
| FVC (z-score) | 228 | -0.1 ($\pm 1.0$) | 259 | 0.2 ($\pm 1.0$) | 65 | 0.4 ($\pm 1.0$) |
| FEV<sub>1</sub>/FVC (z-score) | 228 | -0.6 ($\pm 1.7$) | 259 | -0.4 ($\pm 1.1$) | 65 | -0.6 ($\pm 1.3$) |
| Microbiome samples | 242 |  | 272 |  | 66 |  |
| Nasal |  | 217 (90.4) |  | 272 (100.0) |  | 66 (100.0) |
| Pharyngeal |  | 216 (90.0) |  | 0 (0.0) |  | 0 (0.0) |
| Saliva |  | 240 (99.2) |  | 0 (0.0) |  | 0 (0.0) |

Descriptives are represented by the mean ( ± standard deviation) for continuous variables and the count (proportion in percentage) for categorical variables. \*Asthma exacerbations in the last 12 months. FEV<sub>1</sub>: Forced expiratory volume in 1 second; FVC: Forced vital capacity; BMI: Body mass index.

### Microbiome Epigenome-Wide Association Studies of Exacerbation-Related Bacteria

We tested the association of whole-blood DNAm markers with three α-diversity indices and 18 exacerbation-related bacterial genera, observing low degree of evidence of genomic inflation in any of the mbEWAS performed (λ_GC range:_ 0.97-1.27, **Figures S1-S3**). DNAm levels at 73 CpG sites were significantly associated with the observed bacterial richness and the abundance of five taxa in nasal samples (*Fusobacterium*, *Rothia*, *Streptococcus*, *Neisseriaceae*, and *Leptotrichia*), and with all three α-diversity indices and the abundance of four taxa in pharyngeal samples (*Atopobium*, *Tannerella*, *Campylobacter*, and *Selenomonas*) (FDR<0.05; **Figure 1**, **Table S3**). A total of 20 CpGs surpassed the genome-wide significance threshold (*p*<9×10^-8^; **Table 2**). All associations remained robust in sensitivity analyses for pseudocount variation and for additional microbiome confounders (**Tables S3-S5**). These included genetic loci involved in immune response and inflammation (*TNFAIP8L2*, *LY6G6C*, *CARD14*, *SLIT2*, *PTGDR*, *AOAH*) and insulin secretion and diabetes (*KCNQ1*, *SLC30A5, PTPRN2*). The most significant association in nasal samples was the CpG cg03773820 (*MCPH1*) associated with *Streptococcus* abundance (β=−0.04, *p*=3.7×10^-9^), while in pharyngeal samples it was the CpG cg04739306 (*TFR2*) associated with Shannon diversity (β=0.32, *p*=3.9×10^-10^). No associations at CpG-level were detected for salivary microbiome.

**Figure 1.**
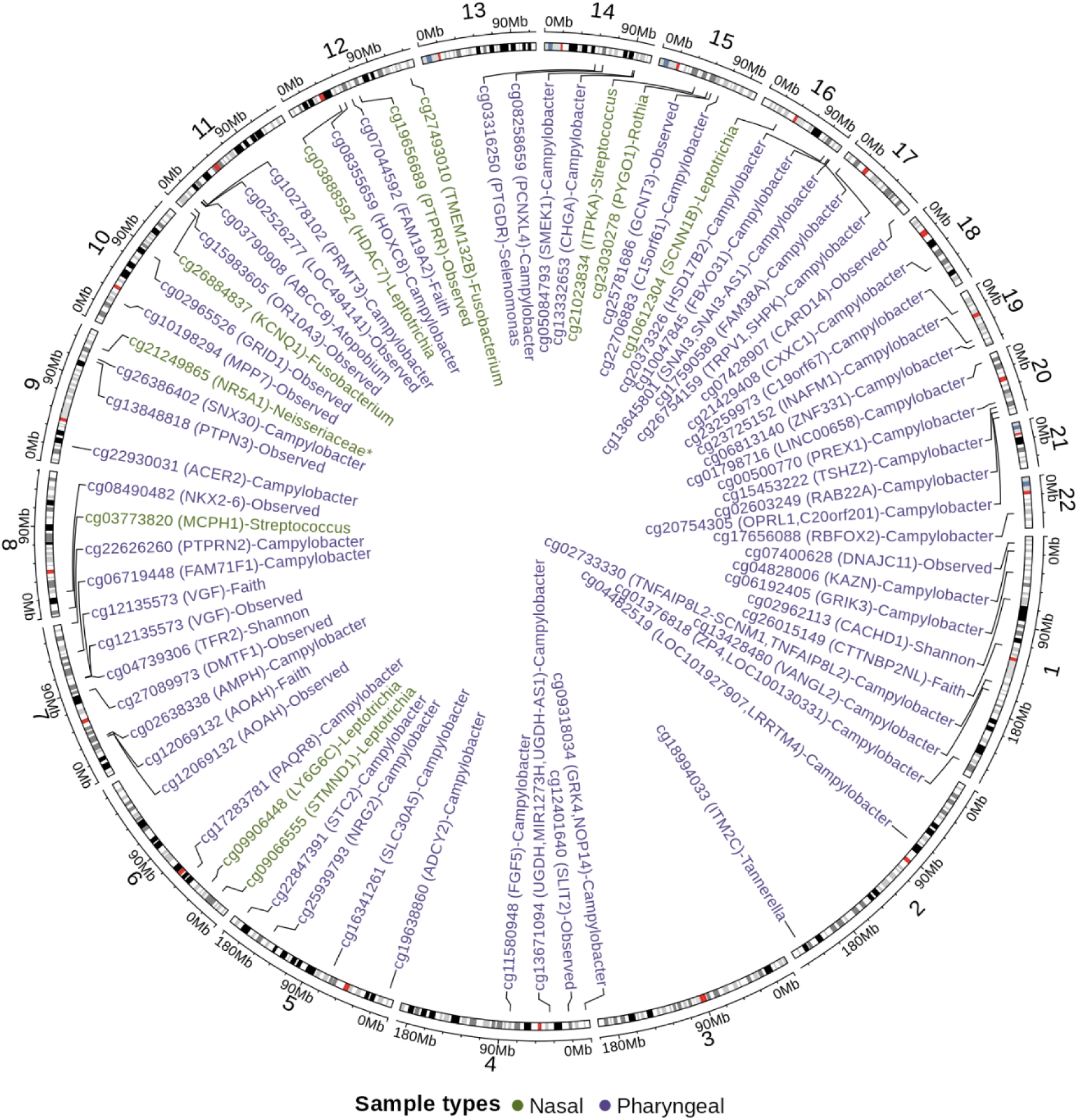
Circos plot representing the 73 CpG sites significantly associated with the upper-airway bacterial genera and diversity indices related to asthma exacerbations. The outer ring represents the human genome (GRCh38/hg38), with chromosomes 1-22 arranged circularly and labeled by number. Cytobands are shown in grayscale, with centromeric regions marked in red. The outer axis indicates chromosomal position in megabases (Mb). Each CpG site is labeled with its probe ID, gene annotation, and the corresponding associated microbiome trait (genus-level taxon or α-diversity index). Labels are color-coded according to sample type: green for nasal samples (n=11) and purple for pharyngeal samples (n=62). *The lowest taxonomy classification available was at the family level. Observed: Observed richness; Shannon: Shannon diversity index; Faith: Faith’s phylogenetic diversity index.

**Table 2.** Summary table of CpGs genome-wide significantly associated with exacerbation-related microbiome traits in GEMAS.

| Sample type | Genus/Index | CpG | Chr | Position* | Gene | $\beta$ | SE | p-value | FDR |
| --- | --- | --- | --- | --- | --- | --- | --- | --- | --- |
| Nasal | *Streptococcus* | cg03773820 | 8 | 6407438 | *MCPH1* | -0.04 | 0.05 | 3.73x10<sup>-9</sup> | 2.59x10<sup>-3</sup> |
|  | Observed | cg19656689 | 12 | 70719749 | *PTPRR* | -0.25 | 0.14 | 3.79x10<sup>-8</sup> | 0.03 |
|  | *Streptococcus* | cg21023834 | 15 | 41493230 | *ITPKA* | -0.05 | 0.06 | 7.57x10<sup>-9</sup> | 2.63x10<sup>-3</sup> |
|  | *Neisseriaceae*<sup>†</sup> | cg21249865 | 9 | 124500073 | *NR5A1* | -0.05 | 0.04 | 2.05x10<sup>-8</sup> | 0.01 |
|  | *Rothia* | cg23030278 | 15 | 55588883 | *PYGO1* | -0.08 | 0.06 | 2.66x10<sup>-8</sup> | 0.02 |
|  | *Fusobacterium* | cg26884837 | 11 | 2542667 | *KCNQ1* | 0.03 | 0.08 | 7.00x10<sup>-8</sup> | 0.02 |
|  | *Leptotrichia* | cg10612304 | 16 | 23317045 | *SCNN1B* | -0.07 | 0.01 | 5.06x10<sup>-8</sup> | 0.02 |
|  | *Leptotrichia* | cg03888592 | 12 | 47817151 | *HDAC7* | -0.04 | 0.01 | 6.51x10<sup>-8</sup> | 0.02 |
|  | *Fusobacterium* | cg27493010 | 12 | 126191121 | *TMEM132B* | -0.07 | 0.01 | 4.68x10<sup>-8</sup> | 0.02 |
| Pharyngeal | Shannon | cg02962113 | 1 | 64686925 | *CACHD1* | 0.20 | 0.23 | 3.24x10<sup>-8</sup> | 0.01 |
|  | *Selenomonas* | cg03316250 | 14 | 52267676 | *PTGDR* | 0.07 | 0.11 | 7.32x10<sup>-10</sup> | 5.09x10<sup>-4</sup> |
|  | *Atopobium* | cg03790908 | 11 | 17476967 | *ABCC8* | 0.07 | 0.09 | 4.53x10<sup>-8</sup> | 0.03 |
|  | Shannon | cg04739306 | 7 | 100633158 | *TFR2* | 0.33 | 0.23 | 3.92x10<sup>-10</sup> | 2.72x10<sup>-4</sup> |
|  | *Campylobacter* | cg04828006 | 1 | 15100034 | *KAZN* | 0.17 | 0.03 | 5.01x10<sup>-9</sup> | 3.48x10<sup>-3</sup> |
|  | *Campylobacter* | cg05084793 | 14 | 91511227 | *SMEK1* | 0.19 | 0.04 | 4.26x10<sup>-8</sup> | 0.01 |
|  | Observed | cg12069132 | 7 | 36545814 | *AOAH* | 0.35 | 0.06 | 4.35x10<sup>-8</sup> | 0.02 |
|  | Observed | cg12135573 | 7 | 101166716 | *VGF* | 0.28 | 0.16 | 2.62x10<sup>-8</sup> | 0.02 |
|  | *Campylobacter* | cg13428480 | 1 | 160400418 | *VANGL2* | -0.14 | 0.12 | 6.23x10<sup>-8</sup> | 0.01 |
|  | *Tannerella* | cg18994033 | 2 | 230868701 | *ITM2C* | -0.20 | 0.12 | 6.99x10<sup>-10</sup> | 4.85x10<sup>-4</sup> |
|  | Faith | cg26015149 | 1 | 112263551 | *CTTNBP2NL* | -1.32 | 1.80 | 8.52x10<sup>-9</sup> | 0.01 |

\*Position based on GRCh38/hg38 build. †The lowest taxonomy classification available was at the family level. Chr: chromosome, SE: standard error, FDR: false discovery rate. Observed: Observed richness; Shannon: Shannon diversity index; Faith: Faith's phylogenetic diversity index.

Among 11 CpG sites associated with the nasal microbiome, the CpG cg19656689 (*PTPRR*) associated with bacterial richness (β=-0.25, *p*=3.8×10^-8^; **Table 2**), was replicated in adolescents from Project Viva (β=-0.07, *p*=0.04; **Table S6**).

### Regional DNAm Analyses

We detected 550 DMRs significantly associated with the exacerbation-related microbiome traits (FDR<0.05; **Figure 2**; **Table S7**), including 160 DMRs identified in nasal, 278 in pharyngeal, and 112 in saliva samples. DMRs included genes involved in allergic airway inflammation (*CHIT1*, *NMUR1*), T-cell activity (*LCP2*, *PIK3CD*, *TXK*, *GNLY*), cytokine and chemokine signaling (*TNFSF10*, *IFNAR1*, *IFITM1*, *N4BP1*), NF-κB (nuclear factor κB) and MAPK (mitogen-activated protein kinase) signaling (*CARD14*, *MAP3K8*, *MAPK15*, *DUSP6*), childhood asthma (*ORMDL3*, 17q12-q21 locus), and obesity and diabetes (*PCSK1*, *MAFA*, *APOB*, *IGF2R*, *SLC2A1*). The top-hit DMRs for each sample type included the gene *AP5Z1* associated with nasal *Neisseria* abundance (five CpGs, *p*=1.3×10^-17^), *SDHA* associated with pharyngeal richness (nine CpGs, *p*=2.2×10^-19^), and *DIRAS3* associated with salivary Faith’s phylogenetic diversity (eight CpGs, *p*=2.7×10^-22^).

**Figure 2.**
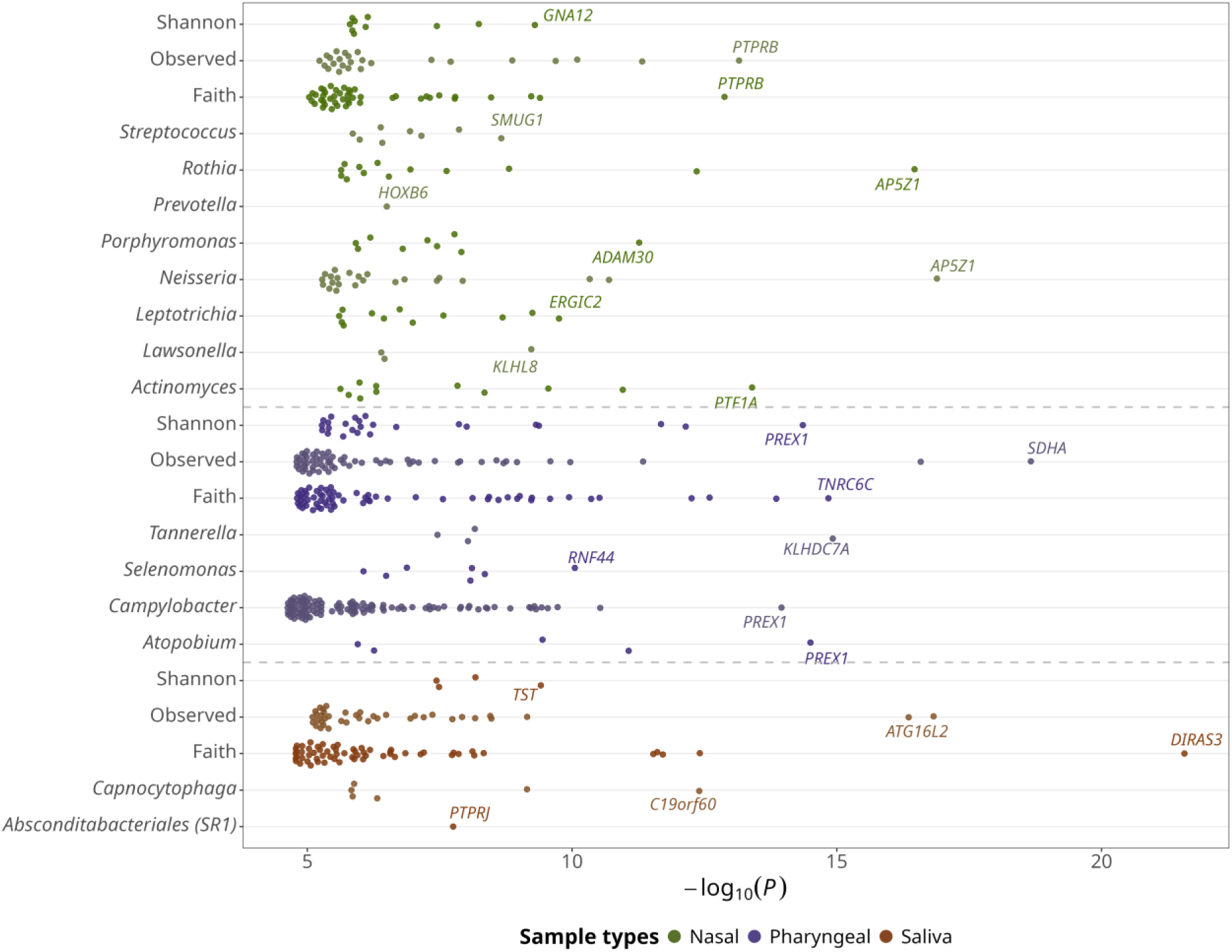
Dot plot showing significant DMRs associated with the α-diversity indices (Shannon’s index, observed richness, and Faith’s phylogenetic diversity) and the bacterial genera previously linked to asthma exacerbations. The x-axis represents the -log_10_(*p*), while the y-axis lists the diversity indices and bacterial genera examined. Each dot represents an individual DMR, colored according to sample type (green: nasal, purple: pharyngeal, brown: saliva), with dashed horizontal lines separating the three sample-type groups. The nearest annotated gene is labeled next to the most significant DMR for each genus or index. Observed: Observed richness; Shannon: Shannon diversity index; Faith: Faith’s phylogenetic diversity index.

Among the 160 DMRs associated with the nasal microbiome and available for replication, five replicated in children with asthma from SysPharmPediA, including associations with Shannon diversity (*GAL3ST2*), *Streptococcus (TEX9)*, *Neisseria (ACAT2)*, and *Rothia (APCDD1L* and *ZBTB38)* (**Table 3**; **Table S8**). In Project Viva, we replicated four DMRs, including *PTPRB* associated with bacterial richness and *Neisseria* and *Rothia* abundances, and *AP5Z1* associated with *Rothia* (**Table 3**; **Table S8**).

**Table 3.**
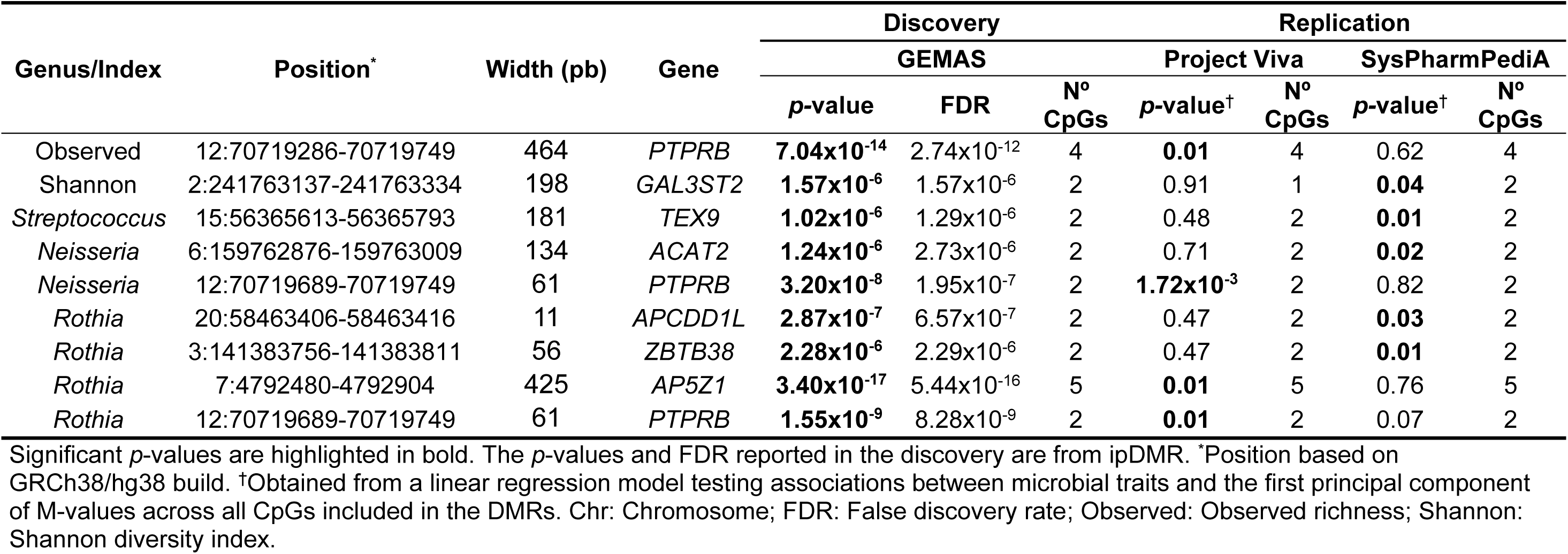
Summary table of the replication for significant DMRs identified in nasal samples.

### Enrichment Analysis

Based on suggestive signals from mbEWAS (*p*<5×10^-5^), we detected an overrepresentation of genes previously associated with childhood-onset asthma, asthma comorbidities and risk factors (e.g., obesity, type 2 diabetes, rheumatoid arthritis, and smoking), inflammatory responses, and the TGF-β (transforming growth factor-β), Wnt, MAPK, and cAMP (cyclic adenosine monophosphate) signaling pathways (**Figure 3**; **Table S9**). We observed enrichment for genes regulated by trichostatin A as well as by the transcription factors NF-κB and the glucocorticoid receptor (*NR3C1*). After stratifying the analyses for saliva, pharyngeal, and nasal samples, we detected sample-specific and shared biological processes (**Table S10**). For example, the MAPK and cAMP signaling pathways were observed in pharyngeal samples, whereas the inflammatory response was primarily observed in nasal samples. In contrast, other terms, such as obesity-related traits, were found in all saliva, nasal, and pharyngeal samples.

**Figure 3.**
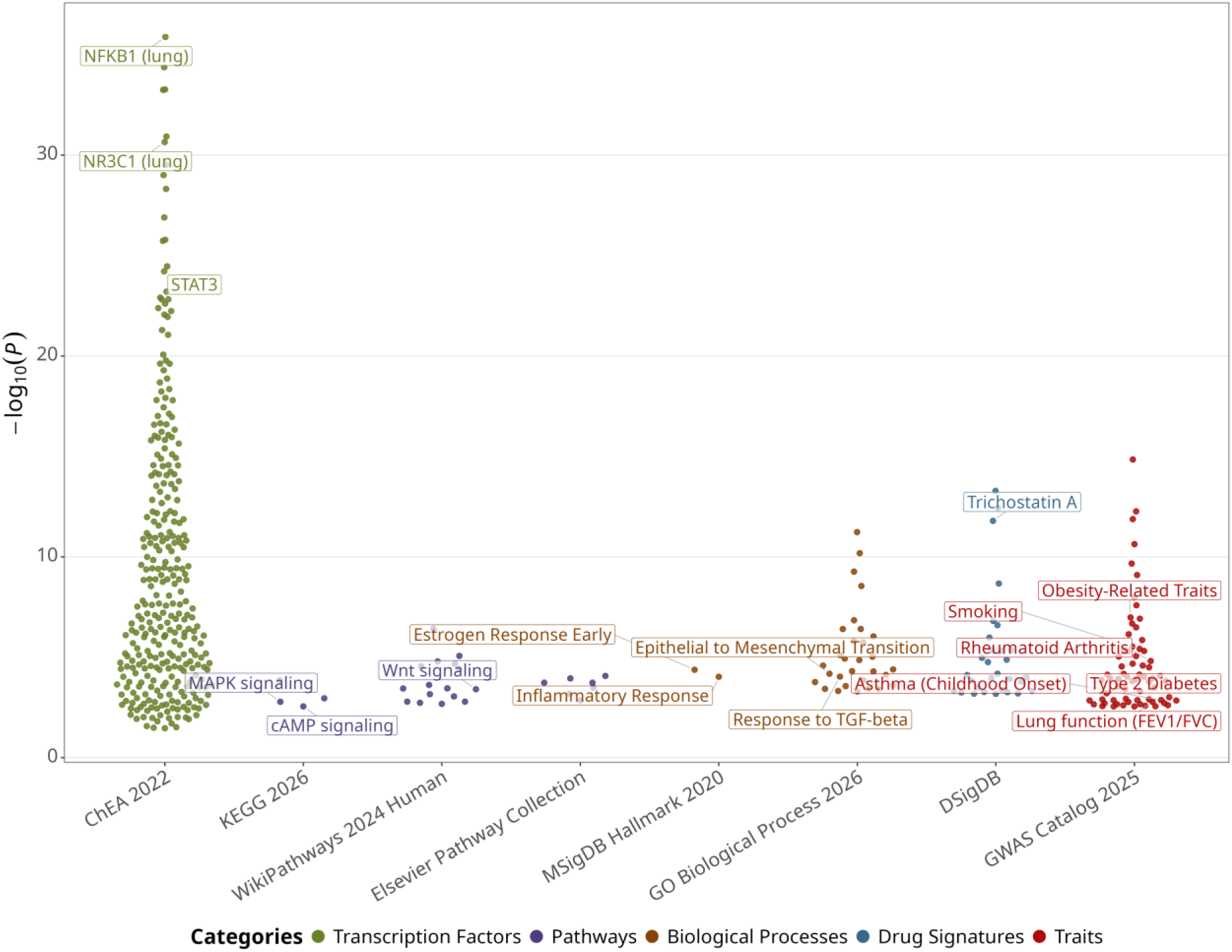
Dot plot of GSEA based on gene annotations from suggestive CpGs identified in the discovery mbEWAS. Each dot corresponds to an enrichment term, which are colored and grouped across the x-axis by category and databases: transcription factors (ChEA 2022), pathways (KEGG 2026, WikiPathways 2024 Human, Elsevier Pathway Collection), biological processes (MSigDB Hallmark 2020, GO Biological Process 2026), drug signatures (DSigDB), and traits (GWAS Catalog 2025). The enrichment -log_10_(*p*) is shown on the y-axis. Relevant terms related to asthma are labeled.

Among nasal enrichment terms, the TGF-β signaling pathway, trichostatin A drug signature, and regulation by NF-κB and *NR3C1* were also observed in SysPharmPediA and Project Viva (*p*<0.05; **Table S11**). Enrichments for genes related to childhood-onset asthma and obesity were also replicated in SysPharmPediA (*p*<0.05).

### Quantitative Trait Loci Analyses

The CpG cg18994033, significantly associated with pharyngeal *Tannerella* abundance, was negatively associated with *ITM2C* blood expression (β=-0.41, FDR=0.03; **Table S12**). This downregulation was *in silico* validated in the eQTM BiosQTL database (*p*=1.6×10^-4^). In addition, four CpGs associated with pharyngeal *Campylobacter* abundance were reported as eQTM in whole blood of *METTL2B*, *HOXC4*, and *MAP2K5* according to the HELIX Project database (*p*_range_: 9.6×10^-5^-1.9×10^-7^; **Table S13**). Furthermore, 43.8% of the CpG sites identified in the mbEWAS (32 out of 73 CpGs) were reported in the EPIC database to be associated with 92 independent meQTLs (FDR<0.05; **Table S14**), including 10 and 82 meQTLs in nasal and pharyngeal bacteria, respectively.

### CpGs Previously Associated with ICS Response or Asthma Exacerbations

Among 22 whole-blood CpGs previously associated with ICS response or asthma exacerbations by EWAS, seven CpGs were associated with exacerbation-related microbiome traits (FDR<0.05; **Table 4**). These included CpGs annotated to *CORT*/*APITD1* associated with nasal *Prevotella*, *FKBP5* and *BAIAP2* associated with pharyngeal *Selenomonas*, *USP7* associated with pharyngeal richness and Faith’s phylogenetic diversity, *C1QL1* and *OTX2* associated with pharyngeal Faith’s phylogenetic diversity, and *OTX2* and *IL12B* associated with salivary *Capnocytophaga* and *Bifidobacterium*, respectively.

**Table 4.** Summary table of significant microbiome associations of CpGs previously related to ICS response or asthma exacerbations.

| Sample type | Genus/Index | CpG | Chr | Position* | Gene | $\beta$ | SE | p-value | FDR |
| --- | --- | --- | --- | --- | --- | --- | --- | --- | --- |
| Nasal | *Prevotella* | cg04256470 | 1 | 10450408 | *CORT, APITD1* | 0.03 | 0.01 | $1.14 \times 10^{-3}$ | 0.02 |
| Pharyngeal | Faith | cg17148519 | 16 | 8899961 | *USP7* | -0.39 | 0.09 | $3.50 \times 10^{-5}$ | $6.99 \times 10^{-4}$ |
| | *Selenomonas* | cg06710464 | 17 | 81073895 | *BAIAP2* | -0.06 | 0.02 | $5.62 \times 10^{-5}$ | $1.12 \times 10^{-3}$ |
| | *Selenomonas* | cg03546163 | 6 | 35686586 | *FKBP5* | 0.12 | 0.04 | $5.34 \times 10^{-4}$ | 0.01 |
| | Observed | cg17148519 | 16 | 8899961 | *USP7* | -0.21 | 0.06 | $6.40 \times 10^{-4}$ | 0.01 |
| | Faith | cg15607672 | 14 | 56810509 | *OTX2* | 0.53 | 0.18 | $3.28 \times 10^{-3}$ | 0.03 |
| | Faith | cg13818573 | 17 | 44968004 | *C1QL1* | 0.24 | 0.09 | $5.97 \times 10^{-3}$ | 0.04 |
| Saliva | *Capnocytophaga* | cg15607672 | 14 | 56810509 | *OTX2* | 0.10 | 0.03 | $1.17 \times 10^{-3}$ | 0.02 |
| | *Bifidobacterium* | cg00066816 | 5 | 159331345 | *IL12B* | 0.05 | 0.02 | $2.10 \times 10^{-3}$ | 0.04 |

\*Position based on GRCh38/hg38 build. Chr: Chromosome; SE: Standard error; FDR: False discovery rate; Faith: Faith's phylogenetic diversity index.

### DNAm Predictors of Plasma Proteins

DNAm-based predicted plasma levels of nine proteins were significantly associated with nasal *Prevotella*, *Streptococcus*, and Faith’s phylogenetic diversity; pharyngeal *Selenomonas*, bacterial richness, and the Shannon index; and salivary *Bifidobacterium* and Faith’s phylogenetic diversity (FDR<0.05; **Figure S4**; **Table S15**). These included proteins involved in immune and inflammatory responses (NOTCH1 and TNFRSF17), tissue remodeling and TGF-β signaling (MMP-9, BMP-1, GDF8), host defense mechanisms (lactoferrin), and glucose metabolism (FGF21).

## DISCUSSION

To the best of our knowledge, this is the first mbEWAS focused on asthma patients, identifying 73 CpGs and 550 DMRs associated with three α-diversity indices and 14 bacterial genera previously related to asthma exacerbations. These included one DMR annotated to *ORDLM3* (17q12-q21 locus), associated with pharyngeal *Campylobacter* abundance, and seven CpGs previously related to ICS response or asthma exacerbations. The CpG cg19656689 (*PTPRR*) and nine DMRs were replicated in independent pediatric populations. The CpG cg18994033, associated with pharyngeal *Tannerella* abundance, was negatively associated with *ITM2C* blood expression. *In silico* meQTL analyses showed that 43.8% of the identified CpGs were likely regulated by genetic variation. We observed an overrepresentation of genes related to asthma comorbidities and risk factors (type 2 diabetes, obesity, smoking), candidate drugs (trichostatin A), transcription factors (NF-κB and glucocorticoid receptor), and biological pathways (TGF-β, Wnt, MAPK, cAMP). Finally, respiratory bacteria were associated with nine DNAm-based predictors of plasma proteins involved in immunoinflammatory responses, tissue remodeling, and TGF-β signaling.

We observed that 64.5% (40/62) of the significant CpGs in pharyngeal samples were associated with *Campylobacter* abundance, including genes involved in airway inflammation (e.g. *TNFAIP8L2*).^23^ This genus has been associated with sputum eosinophil counts and has been correlated with inflammatory mediators increased during asthma exacerbations in children, such as granzyme B, MIP-1β, and PD-L1.^24^ *Leptotrichia* was the taxon with the most significant CpGs in nasal samples (36.4%, 4/11), including the CpG cg03888592 annotated to *HDAC7*, which regulates the expression of Th2 cytokines (e.g., IL-4) and participates in eosinophilic airway inflammation in asthma.^25^ *Leptotrichia* sputum abundance has been associated with asthma and correlated with higher serum IL-4 levels.^26^ Both *Campylobacter* and *Leptotrichia* have shown increased relative abundances in the airways of patients with corticosteroid-resistant asthma.^27^

Among the genome-wide significant CpGs identified in the mbEWAS, several were annotated to genes with evidence for association with asthma, including *PTPRR* (cg19656689, nasal bacterial richness), *PTGDR* (cg03316250, pharyngeal *Selenomonas* abundance), and *AOAH* (cg12069132, pharyngeal bacterial richness). The CpG cg19656689 at *PTPRR* associated with nasal bacterial diversity was replicated in adolescents from Project Viva, supporting its relevance in different age groups. *PTPRR* blood expression is downregulated in severe asthma and implicated in corticosteroid insensitivity through impaired glucocorticoid receptor signaling.^28^ *PTGDR* encodes the prostaglandin D_2_ receptor, a potential therapeutic target for asthma treatment linked to bronchoconstriction, mucosal secretion, and recruitment of T2 inflammatory cells.^29^ *AOAH* genetic polymorphisms have been associated with asthma.^30^ It encodes acyloxyacyl hydrolase, an enzyme that inactivates bacterial lipopolysaccharide (LPS), promotes T2 allergic responses, and increases susceptibility to allergic asthma by preventing persistent LPS-mediated airway epithelial desensitization in mice.^31^

Moreover, we detected one DMR associated with pharyngeal *Campylobacter* abundance annotated to *ORMDL3* (17q12-q21 locus). This is one of the strongest and most consistently replicated genetic susceptibility loci for asthma.^32^ Genetic-microbiome interactions have been described for this gene, where SNPs in *ORMDL3* interact with the upper airway microbiota dominated by *Moraxella*, *Streptococcus*, or *Haemophilus* to increase early-life wheeze risk.^33^ To the best of our knowledge, this is the first study to identify an epigenetic-microbiome interaction involving *ORMDL3*.

We identified that seven CpGs previously associated with ICS response or asthma exacerbation were associated with the exacerbation-related microbiome in our study. Two of these CpGs were annotated to *IL12B* and *FKBP5*, genes with established roles in corticosteroid response. *IL12B* encodes the p40 subunit of IL-12 and IL-23, two cytokines involved in the regulation of immune responses and airway hyperresponsiveness, whose bronchial expression has been associated with corticosteroid responsiveness in asthma.^34^ *FKBP5* encodes a cochaperone that modulates glucocorticoid receptor activity associated with an inflammatory response,^35^ whose bronchial epithelial expression is upregulated by ICS treatment and positively associated with the bronchial microbiome in severe asthma patients.^36,37^ Additionally, we replicated in the SysPharmPediA and Project Viva cohorts an enrichment in genes regulated by the glucocorticoid receptor. Collectively, these findings suggest that microbiome-epigenome interactions may contribute to corticosteroid responsiveness in patients with asthma.

Furthermore, identified DNAm markers, DNAm-based predicted plasma proteins, and enrichment analyses underscored the TGF-β, NF-κB, and MAPK signaling pathways. TGF-β contribute to airway inflammation, tissue remodeling, and fibrosis in asthma,^38^ and enhances bacteria-induced NF-κB-dependent inflammatory responses after lung bacterial infections.^39^ MAPK signaling is key in integrating extracellular signals involved in airway inflammation and remodeling, oxidative stress, and corticosteroid resistance in asthma.^40^ TGF-β, NF-κB, and MAPK pathways are highly interconnected, since activation of TGF-β stimulates NF-κB and MAPK signaling, with the subsequent activation of the Wnt/β-catenin pathway.^41^ Notably, we also observed enrichment of the Wnt signaling pathway, which is highly relevant in asthma, driving processes related to inflammation, airway remodeling, and airway hyperresponsiveness.^42^

Additionally, we observed enrichment of genes implicated in type 2 diabetes and obesity, with the obesity-related enrichment replicated in the SysPharmPediA cohort. Obesity is associated with increased asthma symptom burden, exacerbation frequency, and steroid resistance,^43^ and has been linked to oral and respiratory microbiota dysbiosis involved in obese asthma susceptibility.^44^ In our study, the CpG cg18994033, associated with pharyngeal *Tannerella* abundance, was negatively associated with *ITM2C* blood expression, a gene differentially expressed in Th cells from children with obesity-related asthma.^45^ Diabetes mellitus is also associated with severe asthma phenotypes, greater risk of emergency room visits, and hospitalizations.^46^ Although the mechanisms linking obesity and type 2 diabetes to asthma remain unclear, these have in common systemic inflammation^44,46^ and bacterial dysbiosis^27,44,47^. Moreover, we observed and replicated in SysPharmPediA and Project Viva an enrichment of genes regulated by trichostatin A, a histone deacetylase inhibitor and a candidate drug for asthma, diabetes, and obesity.^48,49^

We found consistencies between these epigenetic findings and genetic signals from our previous mbGWAS, focused on the same exacerbation-related upper-airway microbiome.^9^ Using genetic and epigenetic approaches, we observed enrichment for genes related to obesity and smoking and regulated by trichostatin A, NF-κB, and the glucocorticoid receptor.^9^ This supports the relevance of host-microbiome interactions in asthma comorbidities and asthma exacerbations treatment through genetic and epigenetic mechanisms. Our results are also consistent with the two previous mbEWAS of the respiratory microbiome. Bozack et al. also identified microbiome-associated CpGs annotated to genes involved in asthma, inflammation, and immune function^11^ although they did not report any specific CpG of DMR overlapping with our study. Shibata et al. linked *Streptococcus pneumoniae*-associated DMRs to serum MMP-9 in infants with severe bronchiolitis, a protein elevated in bronchoalveolar lavage fluid from adults with severe asthma.^10^ Similarly, we found that DNAm-predicted plasma MMP-9 levels were associated with *Prevotella* abundance, with a trend also for *Streptococcus* (FDR=0.07).

This study has several strengths. First, we performed the first mbEWAS focused on patients with asthma by integrating microbiome data from three asthma-relevant upper-airway samples with blood DNAm profiles. Second, we provided replication in two independent cohorts. Third, we applied state-of-the-art methods to analyze microbiome and epigenome data, including adjustment for potential confounders, correction for multiple testing, and sensitivity analyses. Fourth, we used omics approaches without prior hypotheses, enabling the discovery of novel biomarkers.

Nevertheless, some limitations should be acknowledged. First, the cross-sectional design does not allow causal and temporal inference. Second, replication may be limited by the heterogeneity among analyzed populations and the unavailability of pharyngeal and saliva microbiome data in replication cohorts. While most of the participants from the discovery phase were adults, all participants in the replication phase were children, which likely present different asthma phenotypes with distinct molecular signatures.^50^ Third, microarrays covered only ∼3% of CpG sites across the human epigenome,^51^ leaving the remaining epigenetic markers unexplored. Fourth, the targeted sequencing of the 16S rRNA gene does not provide species-level resolution and excludes other relevant microorganisms beyond bacteria, which will be essential for completely understanding the microbiome-epigenome crosstalk.

## CONCLUSION

We identified novel associations between whole-blood DNAm markers and the upper-airway bacterial genera and diversity indices associated with asthma exacerbations despite ICS treatment. Identified epigenetic loci were mainly involved in immune and inflammatory responses, including *ORMDL3* at the 17q12-q21 asthma susceptibility locus and a replicated signal at *PTPRR* implicated in corticosteroid insensitivity. We observed an overrepresentation of genes involved in asthma-related comorbidities, biological pathways, and candidate therapeutic targets. Additionally, we detected associations of the exacerbation-related microbiome with DNAm-based predicted plasma proteins involved in immunoinflammatory responses, tissue remodeling, and TGF-β signaling. These findings support a role for microbiome-epigenome interactions in asthma exacerbations that should be further explored to advance precision medicine in asthma.

## AUTHOR CONTRIBUTIONS

F.L.-D., M.P.-Y. and J.P.-G. contributed to the conceptualization of the study; E.C.-V., J.P.-G., and M.P.-Y. to the methodology; E.C.-V., M.M.-A., E.L.G., E.M.-G., F.L.-D., and J.P.-G. to formal analysis and investigation; R.G.-P., J.M.H.-P., O.S.-P., P.P.-G., E.M.-L., P.C.-E., I.S.-M., L.L.-F., S.L.R.-S., S. Hashimoto, S.J.H.V., A.H.N., H.B.-D., M.G., A.A.T., S. Harner, S.B., C.W., U.P., M.K., A.D.K., J.V., M.-F.H., E.O., D.R.G., M.I.A.-A., A.H.M., A.C., F.L.-D., and M.P.-Y. to resources, cohort management, and/or patient recruitment; E.C.-V., M.M.-A., A.B., E.L.G., E.M.-G., and J.P.-G. to data curation; E.C.-V. to visualization; E.C.-V. and J.P.-G. to data interpretation and writing of the original draft; M.P.-Y. and J.P.-G. to funding acquisition and project supervision. All authors contributed to the critical revision and editing of the manuscript, approved the final version, and agreed to be accountable for all aspects of the work, ensuring its accuracy, integrity, and scientific rigor.

## DATA AVAILABILITY

All summary statistics supporting the findings and conclusions presented in this manuscript are provided in the main text, Online Supplement, and/or deposited in the Zenodo repository (accession number 10.5281/zenodo.23036316). The Zenodo dataset will be made publicly accessible upon manuscript acceptance and available to reviewers during the peer-review process. Summary statistics for significant CpG sites identified in the discovery mbEWAS will also be made publicly available through the EWAS Atlas following manuscript acceptance. 16S-rRNA sequences from the GEMAS study are available in the Sequence Read Archive under accession number PRJNA878647. Individual-level demographic and epigenomic data from the analyzed cohorts cannot be made publicly available due to ethical restrictions and the consent provided by participants. Data access may nevertheless be considered upon reasonable request to the corresponding author, subject to approval by principal investigators of the studies and ethics committees.

## ETHICAL APPROVAL

The GEMAS study has been approved by the ethics committees of the participating centers (approval 29/17 for the hospitals in the Canary Islands and PI2019077 for the Hospital Universitario Donostia). The SysPharmPediA consortium has also been approved by the ethics committees of the participating centers (approval numbers: 18-1034-101 for centers in Germany; PI2015075[SO] in Spain; NL55788.041.15 in the Netherlands; 0120-569/2017/4 in Slovenia; Dnr 2010/1336-31/3 and 2019-00546 in Sweden). All protocols from the pre-birth Project Viva cohort were approved by the Harvard Pilgrim Health Care Institutional Review Board.

## ACKNOWLEDGMENTS

We gratefully acknowledge the patients, families, recruiters, healthcare providers, and community clinics for their participation in the GEMAS, SysPharmPediA, and Project Viva studies. In addition, the author(s) wish to acknowledge the contribution of the Teide High-Performance Computing (TeideHPC) facilities to the results of this research. TeideHPC is part of the Atlantic Supercomputing Centre (Centro de Supercomputación del Atlántico) at the Instituto Tecnológico y de Energías Renovables (ITER, SA). URL: http://teidehpc.iter.es.

## AI STATEMENT

ChatGPT (OpenAI) and Claude (Anthropic) were used to improve the English language, grammar, and readability of the manuscript, and to assist in writing and optimizing code. All AI-assisted text and code were reviewed and validated by the lead and corresponding authors, and all authors take full responsibility for the content of this article.

