## Supplementary Methods for "Epigenome-Wide Association Study of the Upper-Airway Microbiome Involved in Asthma Exacerbations"

#### Design and Study Populations

We followed a discovery and replication study design, in which three cohorts from Europe and the United States were analyzed independently. The discovery phase included participants from the GEMAS study recruited in Spain, while the replication phase included the SysPharmPediA and Project Viva cohorts. All studies were approved by the corresponding ethics committees of the participating centers and were conducted in accordance with the International Code of Medical Ethics of the World Medical Association (Declaration of Helsinki). All participants provided written informed consent. For those aged <18 years old, written consent from legal guardians and written assent from the participants were obtained. In all cohorts, demographic and clinically relevant asthma-related variables were recorded using standardized questionnaires. A description of each cohort is provided below.

*Genomics and Metagenomics of Asthma Severity (GEMAS)*: Ongoing observational study aimed at characterizing the influence of microbiome and human genetic variation on asthma exacerbations (ClinicalTrials.gov ID: NCT04501926).^S1^ Briefly, asthma patients were recruited between 2018 and 2020 from Allergy and Pulmonology departments in hospitals across the Canary Islands (Hospital Universitario de Canarias, Hospital Universitario N.S. de Candelaria, and Hospital Universitario de La Palma) and the Basque Country (University Hospital Donostia). Inclusion criteria were: (1) male and female participants aged 8-85 years old, (2) asthma diagnosis according to the Global Initiative for Asthma (GINA) 2020 guidelines,^S2^ and (3) ongoing asthma treatment corresponding to GINA therapeutic steps 1-5. Patients were excluded if they met any of the following conditions: (1) at least one grandparent of non-European origin, (2) pregnancy, (3) coexistence of other chronic respiratory diseases, or (4) known first- or second-degree kinship with other participants enrolled in the study. In the present work, a subset of 242 patients was analyzed, comprising 191 mostly adults from the Canary Islands and 51 children from the Basque Country, for whom saliva, nasal, and pharyngeal samples were available for microbiome analyses and blood samples for epigenomics.

*Systems Pharmacology Approach to Uncontrolled Pediatric Asthma (SysPharmPediA)*: Multicenter and multiethnic consortium that recruited children with moderate-to-severe asthma in four European countries (Spain, the Netherlands, Slovenia, and Germany) (ClinicalTrials.gov ID: NCT04865575).^S3^ Briefly, the inclusion criteria were: (1) children aged 6-17 years, (2) a clinical diagnosis of asthma, and (3) moderate to severe asthma treated with a therapeutic step ≥3 according to GINA guidelines.^S4^ The analysis included 66 individuals with available blood and nasal samples for epigenomic and microbiome analyses, respectively.

*Project Viva*: Longitudinal pre-birth cohort that initially recruited pregnant women between 1999 and 2002 during their first prenatal visits at Atrius Harvard Vanguard Medical Associates (Massachusetts, USA). A complete description of the cohort can be found elsewhere.^S5,S6^ Eligibility criteria included: (1) English-speaking mothers, (2) gestational age <22 weeks at enrollment, and (3) a singleton pregnancy. Of the original birth cohort, 574 children were recontacted during early adolescence (13 years of age) and provided informed consent/assent for the collection of nasal and blood samples for epigenomic and microbiome analyses. We analyzed a subset of 272 participants with blood DNAm and nasal microbiome data, after excluding related participants and those with missing data for asthma, exacerbations, and covariates.

#### Assessment of Clinical Variables

Demographic and clinical variables were collected in GEMAS by physicians and health care recruiters using standardized questionnaires, as described elsewhere.^S1^ Body mass index (BMI) was categorized as normal weight, overweight, or obese using BMI cutoffs (≥19 years) or BMI-for-age z-scores (<19 years) based on the World Health Organization (WHO) guidelines.^S7^ Lung function parameters were expressed as z-scores using the race-neutral Global Lung Function Initiative (GLI) 2022 reference equations.^S8^ Asthma exacerbations in the past year were defined as the need for systemic corticosteroid use, hospitalization, and/or emergency room visits due to asthma.^S9^

#### Microbiome Profiling in the Discovery Cohort

The nasal, pharyngeal, and saliva microbiome were profiled by targeted sequencing of the 16S rRNA gene (V3-V4 region) following Illumina’s official guidelines (Illumina, San Diego, CA) and protocols adapted for microbiome studies.^S10^ A comprehensive description of the methodology, including library preparation, sequencing, and downstream data processing, has been reported elsewhere.^S11^ Briefly, we employed microbiome-specific kits for sample collection and processing, assayed positive and negative controls, denoised the sequencing reads and removed contaminant and non-bacterial reads, and filtered out samples with <10,000 reads. After QC, 222 nasal, 224 pharyngeal, and 249 saliva samples were retained from an initial set of 263 samples in the GEMAS cohort.

Following the MiBioGen consortium guidelines,^S12^ we focused on the bacterial genera present in at least 10% of samples. Read counts were rarefied to 10 000 reads per sample to normalize sequencing depth, and data were log-transformed to reduce skewness and the influence of highly abundant taxa. Before log-transformation, we added a pseudocount of 1 due to the presence of zeros. In addition, α-diversity indices were calculated from the rarefied data at the ASV level.^S13^ Three α-diversity indices were computed: Observed richness, Shannon index, and Faith’s phylogenetic diversity. Observed richness and Faith’s phylogenetic diversity were also log-transformed, whereas the Shannon index remained unchanged, as it is inherently based on a logarithmic formulation. In the GEMAS Basque Country subgroup, the genera *Dialister* (nasal samples) and *Bifidobacterium* (saliva samples) were excluded as they were present in <10% of samples.

#### Microbiome Profiling in the Replication Cohorts

In the SysPharmPediA consortium, nasal microbiome data were generated and processed following the same procedures as those applied in the discovery cohort. Briefly, paired-end reads (2x300 bp) were generated on the MiSeq platform (Illumina). To ensure data quality, a simulated mock community was included as a positive control, and negative controls, together with DNA extraction and PCR replicates, were sequenced throughout the workflow.

Demultiplexed 16S rRNA sequencing reads were analyzed using the bioinformatic tool QIIME2.^S14^ Low-quality bases were trimmed according to their Phred scores, and reads then underwent quality control (QC) (i.e., denoising) and were resolved into amplicon sequence variants (ASVs) using the DADA2 algorithm.^S15^ To remove non-bacterial sequences, reads were aligned against the SILVA rRNA v138 reference database^S16^. For taxonomic assignment, a Naïve Bayes classifier was trained specifically for the V3-V4 region of the 16S rRNA gene using the SILVA v138 database. ASVs were aligned using MAFFT,^S17^ and rooted phylogenetic trees were inferred with FastTree2^S18^ for subsequent calculation of Faith’s phylogenetic diversity index. Following QIIME2 processing, data were imported into R (v4.4.2) and managed with the *phyloseq* package.^S19^ Contaminant ASVs were identified and removed using the *decontam* R package.^S20^ Additionally, samples with <10,000 high-quality reads were discarded. After QC, 81 nasal samples were retained from an initial set of 98 samples. Additionally, the genus *Dialister* was excluded from the analyses because it was present in <10% of samples.

In the Project Viva cohort, a complete description of the microbiome profiling methodology is available elsewhere.^S21^ In this cohort, the nasal microbiome was profiled by targeted sequencing of the V1–V3 hypervariable regions of the 16S rRNA gene. Sequencing reads were clustered into operational taxonomic units (OTUs), and contaminants, low-abundance OTUs, and samples with fewer than 10,000 reads were removed during QC. Following QC, 416 of the 436 initial samples were retained for subsequent analyses. In addition, the genera *Dialister* and *Lawsonella* were excluded because they were present in <10% of samples, and Faith’s phylogenetic diversity index was not calculated since ASV resolution was not available.

#### Processing of Epigenomic Data

Epigenomic data were generated in GEMAS from genomic DNA extracted from blood samples using Wizard Genomic DNA Purification Kits (Promega, Fitchburg, WI). The Infinium MethylationEPIC v1.0. BeadChip (Illumina) was used to interrogate DNAm levels at 865,918 CpG sites across the genome. A complete protocol of the data processing pipeline is provided elsewhere.^S22^ Briefly, CpG sites were excluded if they showed (1) poor-quality measurements in >5% of individuals, (2) missing data in >5% of individuals, (3) multimodal distributions, (4) annotation to sex chromosomes, (5) non-specific alignment to multiple genomic regions, and/or (6) evidence of genetic variation at the methylation probe (polymorphic probes). Polymorphic probes were defined based on the presence of single nucleotide polymorphisms with a minor allele frequency >1% based on Illumina manifests. Samples were excluded if they presented (1) poor-quality data in >5% of CpG sites, (2) discordance between reported and methylation-predicted sex, (3) missing data in >10% of CpG sites, and/or (4) atypical signal distributions in genotype control probes inconsistent with expected homozygous or heterozygous patterns. After QC, 703,505 CpG sites and 677 samples were retained. Among these, 242 samples had microbiome data available. β-values were log-transformed to M-values to minimize skewed distributions,^S23^ and all analyses were conducted on M-values.

In the SysPharmPediA consortium, genomic DNA from blood samples was isolated using the FlexiGene DNA Kit (Qiagen) and DNAm levels were measured using the Infinium MethylationEPIC v1.0 BeadChip (Illumina). QC was conducted using the same procedures applied in the discovery cohort, as described elsewhere.^S24^ From an initial set of 131 samples, 127 were retained after QC, of which 66 had available microbiome data.

In the Project Viva cohort, genomic DNA was extracted from blood samples, and DNAm was measured using the Infinium MethylationEPIC v1.0 BeadChip (Illumina). The data processing workflow has been described in detail elsewhere.^S25^ Following QC, 654 of 688 samples were retained. Among these, 272 had available microbiome data.

Batch effects in DNAm data were corrected in GEMAS and Project Viva studies using the *ComBat-met* R package adjusting for age, sex, and ancestry (race and ethnicity in Project Viva due to lack of genotype data).^S26^ All SysPharmPediA samples were run in the same batch for epigenetic analyses.

#### Microbiome Epigenome-Wide Association Studies

Microbiome epigenome-wide association studies (mbEWAS) were conducted in the discovery population to identify blood DNAm markers associated with the upper airway microbiome, specifically with three α-diversity indices (observed richness, Shannon’s index, and Faith’s phylogenetic diversity) and 18 bacterial genera previously related to asthma exacerbations despite ICS treatment in GEMAS.^S11^ The selected genera were *Porphyromonas*, *Prevotella*, *Streptococcus*, *Fusobacterium*, *Lawsonella*, *Dialister*, *Neisseriaceae* spp., *Neisseria*, *Leptotrichia*, *Actinomyces*, and *Rothia* from nasal samples; *Selenomonas*, *Atopobium*, *Tannerella*, and *Campylobacter* from pharyngeal samples; and *Absconditabacteriales*, *Bifidobacterium*, and *Capnocytophaga* from saliva samples. We also analyzed the three α-diversity indices assessed in each of the three sample types.

The association between blood DNAm and bacterial abundance or diversity was assessed using multivariate linear regression models implemented in the *limma* R package.^S27^ In the discovery cohort, models were adjusted for age, sex, genetic ancestry, cell-type heterogeneity, and microbiome sequencing pool. We used the reference-free ReFACTor algorithm for correction of cell heterogeneity,^S28^ selecting six and four ReFACTor principal components (PCs) in the Canary Islands and Basque Country subgroups, respectively. Genetic ancestry was estimated from genotype data (generated using the Illumina Global Screening Array v3.0), as described elsewhere,^S29^ and the first two PCs were selected for adjustment. mbEWAS were performed separately within each subgroup due to age differences, followed by a meta-analysis using the software METASOFT.^S30^ Based on Cochran’s Q *p*-value, a fixed-effect model was applied for CpGs with low heterogeneity across studies (*p*>0.05), and a random-effect model for those with high heterogeneity (*p<*0.05). As in the GEMAS Basque Country subgroup the genera *Dialister* and *Bifidobacterium* were excluded, results for these taxa were not meta-analyzed and are based solely on the GEMAS Canary Islands subgroup.

Genomic inflation was evaluated through quantile-quantile (Q-Q) plots and estimation of the genomic inflation factor (λ_GC_, **Figures S1-S3**). Multiple testing was corrected for each bacterial genus or diversity index using a false discovery rate (FDR)<0.05 based on the Benjamini–Hochberg procedure, and genome-wide significance was declared at *p*<9x10^-8^, as calculated for EPIC v1.0 array.^S31^ Significant CpG sites without a gene annotation based on the Illumina manifest file v1.0 B5 were annotated to the nearest gene using the *rGREAT* R package.^S32^

**Sensitivity analyses**

To evaluate whether the results were dependent on the choice of pseudocount, the default pseudocount of 1 was replaced with alternative values of 0.5 and 0.1, and all mbEWAS were repeated under both settings. Associations identified in the discovery analysis were reported only if they remained significant (p<0.05) using both alternative pseudocounts. In addition, sensitivity analyses were conducted to assess the robustness of the significant findings to potential confounders of microbial communities previously identified in the GEMAS study.^S11^ For nasal samples, these confounders included antibiotic prescription in the last two months, BMI, season of sampling, and smoking within 30 minutes prior to sample collection, whereas for pharyngeal samples they included antibiotic prescription in the last two months, cavities, and liquid intake within 30 minutes prior to sample collection.

#### Differentially Methylated Regions

Based on the mbEWAS results, differentially methylated regions (DMRs) were identified using two independent methods: ipDMR^S33^ and comb-p^S34^ from the *Enmix* R package.^S35^ DMRs were estimated by aggregating CpG sites located within a 1 kb window. Only regions containing at least 2 CpGs that reached regional significance (FDR<0.05) in both tools were retained. To ensure consistency, only DMRs showing >50% overlap of the genomic coordinates between ipDMR and comb-p results were retained. Identified DMRs were annotated to the nearest gene using the *rGREAT* package.^S32^

#### Enrichment Analyses

Gene-set enrichment analyses (GSEA) were performed to identify biological processes, human diseases and traits, and drug-related genetic signatures overrepresented among gene annotations of CpG sites identified in the nasal, pharyngeal, and saliva discovery mbEWAS. A suggestive significance threshold of *p*<5x10^-5^ was used for CpG selection. Genes were grouped into one dataset combining the mbEWAS from the three biological samples. GSEA was conducted using the tool Enrichr^S36^ which allows querying multiple curated databases. Results with FDR<0.05 were considered statistically significant. To assess robustness, enrichment analyses were repeated using alternative CpG-selection thresholds (*p*<5x10^-4^ and *p*<5x10^-6^). Findings were reported only if they remained significant across both thresholds (*p*<0.05). To evaluate the contribution of each biological sample type, stratified enrichment analyses were also performed separately for nasal, pharyngeal, and saliva-derived gene sets.

The database GWAS Catalog 2025 was used to assess enrichment in phenotypes and human diseases, the Drug Signatures Database (DSigDB) for drug-related genetic signatures, Gene Ontology Biological Process 2026 and Molecular Signature Database Hallmark (MSigDB 2020) for biological processes, the ChIP Enrichment Analysis (ChEA 2022) database for transcription factor binding sites, and the KEGG 2026, WikiPathways 2024 Human, and Elsevier Pathway Collection, for pathways.

#### Replication analyses

We only followed up for replication the discoveries from the nasal microbiome given the availability in replication datasets. We followed up for replication CpGs, DMRs, and enrichment terms identified in the discovery phase at FDR<0.05 and robust to sensitivity analyses.

Replication of CpGs

Associations between whole-blood DNAm at CpGs and microbiome traits were evaluated using linear regression models in *limma*, as in the discovery phase. In SysPharmPediA, mbEWAS models were adjusted for age, sex, genetic ancestry (six genotype PCs), cell-type heterogeneity (six ReFACTor PCs), and microbiome sequencing pool. In Project Viva, models were adjusted for age, sex, asthma status, self‐reported race and ethnicity, cell-type heterogeneity (five ReFACTor PCs), and microbiome sequencing pool. Replication was declared in each cohort for significant associations (*p<*0.05) showing the same direction of effect as in the discovery cohort.

Replication of DMRs

For each DMR identified in the discovery, DNAm levels were summarized using the first principal component (PC_1_) derived from the M-values of all CpGs located within the corresponding genomic region. PC_1_ was used as a proxy for the overall methylation level of the DMR. Associations between DMR methylation and microbiome traits were then assessed using multivariate linear regression models, with PC_1_ as the dependent variable, the microbiome trait (genus abundance or α-diversity index) as the independent variable, and the same covariates included in the discovery mbEWAS. The same pipeline was applied in SysPharmPediA and Project Viva cohorts, and replication was declared for associations reaching significance (*p*<0.05) and showing the same direction of effect as in the discovery cohort. In addition, each DMR was required to have at least two CpGs available in the replication cohorts to be considered replicated.

Replication of enrichment terms

GSEA in the replication cohorts were performed using the same procedure applied in the GEMAS study. Only enrichment terms from nasal samples that remained robust to variations in the CpG selection threshold during the discovery phase were taken forward for replication. Terms reaching statistical significance (*p*<0.05) in the corresponding replication cohort were considered replicated.

#### Expression Quantitative Trait Methylation Analyses

Whole-blood RNA sequencing (RNA-seq) data were obtained from participants of the discovery cohort (GEMAS), including both asthma patients and non-asthmatic controls from the Canary Islands. Whole-blood samples of 119 participants were collected in PAXgene Blood RNA tubes (Qiagen), and total RNA was extracted using the NucleoSpin® RNA Blood kit (Macherey-Nagel, Düren, Germany) following the manufacturer’s instructions. To avoid the overrepresentation of hemoglobin transcripts, globin mRNA was depleted from the total RNA using the GlobinClear™ Human Whole Blood Globin Reduction Kit (Thermo Fisher Scientific/Invitrogen, Carlsbad, CA) according to the manufacturer's protocol. RNA-seq libraries were subsequently prepared using the NGS RNA Library Prep Set (PT042; Novogene, Beijing, China) as recommended by the manufacturer. Libraries were quantified, pooled, and sequenced on a NovaSeq X Plus Series to generate 150 bp paired-end reads. Raw sequencing reads were subjected to quality control using FastQC.^S37^ High-quality reads were then aligned to the GRCh38 human reference genome using STAR^S38^ and GENCODE v30 gene annotations. Gene-level expression was quantified with RSEM.^S39^ Gene-count data were imported into R using *tximport*.^S40^ Genes with zero length and zero variance across samples were removed, and lowly expressed genes were subsequently filtered using the *filterByExpr* function from the *edgeR* package,^S41^ retaining 19,699 genes for downstream analyses. Finally, gene counts were normalized using the trimmed mean of M-values (TMM) method and transformed by rank-based inverse normal transformation using the *RNOmni* package.^S42^

In the eQTM analysis, to account for unobserved technical and biological sources of variation, surrogate variable (SV) analysis was performed using the *sva* package.^S43^ SVs were estimated with the *svaseq* function, adjusting the null model for age, sex, genetic ancestry (first two genotype PCs), and the first five cell-composition components estimated from DNA methylation data using ReFACTor.^S28^ The full model additionally included asthma status and 12 significant SVs were identified. To minimize collinearity and avoid overadjustment, correlations between SVs and ReFACTor components were evaluated. SVs showing significant correlations with any ReFACTor component (FDR<0.05 and |r|>0.5) were excluded from subsequent analyses. Consequently, three SVs were removed, and the remaining nine SVs were included as covariates.

The eQTM analysis focused on CpG sites identified in the discovery mbEWAS as significantly associated with upper airway microbiome features related to asthma exacerbations (FDR<0.05). Associations between DNA methylation M-values at these CpGs and the expression of nearby genes were evaluated, restricting analyses to cis pairs defined as genes with a transcription start site located within 100 kb of the CpG site. eQTM analyses were performed using the *MatrixEQTL* package under a linear regression framework,^S44^ adjusting for age, sex, asthma status, the first two genotype PCs, the first five ReFACTor components, and the nine retained SVs. Associations with an FDR<0.05 were considered statistically significant.

#### *In Silico* Quantitative Trait Analyses

The associations between the identified significant CpGs from the mbEWAS and genetic variation (meQTL) or gene expression (eQTM) were also evaluated using publicly available databases. Specifically, MeQTL EPIC^S45^ and BIOS QTL^S46^ were used for meQTL identification, while the HELIX Project^S47^ and BIOS QTL were employed for eQTM detection. The MeQTL EPIC database contains meQTL results for 724,499 CpGs profiled with the Infinium MethylationEPIC (Illumina) array in 2,358 blood samples from three UK cohorts.^S45^ The HELIX Project provides eQTM results for 386,518 CpGs measured with the Infinium HumanMethylation450K (Illumina) array in 832 blood samples from children of European ancestry.^S47^ BIOS QTL includes both meQTL and eQTM results for 405,709 CpGs profiled with the Infinium HumanMethylation450K (Illumina) array in 3,841 and 2,101 blood samples, respectively, from five Dutch biobanks.^S46^

#### Assessment of CpGs Previously Related to ICS Response or Asthma Exacerbations

As a candidate approach, we investigated whether CpGs previously associated with ICS response or asthma exacerbations by EWAS until June 2026 were also associated with the upper airway microbiome traits linked to asthma exacerbations.^S48^ A total of 22 CpGs associated with ICS response or asthma exacerbations were included in the analysis (**Table S2**). Multiple testing was controlled using an FDR<0.05, considering the number of candidates CpGs evaluated.

#### DNAm Predictors of Plasma Proteins

DNAm-based predictors were derived using the *methscore* function implemented in the *ENmix* package,^S49^ which included predictors of 109 plasma protein levels. Associations between each DNAm predictor and the upper airway microbiome traits linked to asthma exacerbations were tested in GEMAS using multivariate linear regression models adjusted for age, sex, genetic ancestry, cell-type heterogeneity, and microbiome sequencing pool. Analyses were performed separately within each GEMAS subgroup (Canary Island and Basque Country), and results were meta-analyzed using METASOFT.^S30^ Statistical significance was determined using an FDR<0.05, considering the number of plasma proteins evaluated.

S37.	Andrews S. FastQC: A Quality Control Tool for High Throughput Sequence Data. http://www.bioinformatics.babraham.ac.uk/projects/fastqc

S38.	Dobin A, Davis CA, Schlesinger F, Drenkow J, Zaleski C, Jha S, et al. STAR: ultrafast universal RNA-seq aligner. *Bioinformatics*. 2013;29(1):15-21. doi:10.1093/bioinformatics/bts635

S39.	Li B, Dewey CN. RSEM: accurate transcript quantification from RNA-Seq data with or without a reference genome. *BMC Bioinformatics*. 2011;12(1):323. doi:10.1186/1471-2105-12-323

S40.	Soneson C, Love MI, Robinson MD. Differential analyses for RNA-seq: transcript-level estimates improve gene-level inferences. *F1000Research*. 2016;4:1521. doi:10.12688/f1000research.7563.2

S41.	Chen Y, Chen L, Lun ATL, Baldoni PL, Smyth GK. edgeR v4: powerful differential analysis of sequencing data with expanded functionality and improved support for small counts and larger datasets. *Nucleic Acids Res*. 2025;53(2):gkaf018. doi:10.1093/nar/gkaf018

S42.	McCaw ZR, Lane JM, Saxena R, Redline S, Lin X. Operating characteristics of the rank‐based inverse normal transformation for quantitative trait analysis in genome‐wide association studies. *Biometrics*. 2020;76(4):1262-1272. doi:10.1111/biom.13214

S43.	Leek JT, Johnson WE, Parker HS, Jaffe AE, Storey JD. The sva package for removing batch effects and other unwanted variation in high-throughput experiments. *Bioinformatics*. 2012;28(6):882-883. doi:10.1093/bioinformatics/bts034

S44.	Shabalin AA. Matrix eQTL: ultra fast eQTL analysis via large matrix operations. *Bioinformatics*. 2012;28(10):1353-1358. doi:10.1093/bioinformatics/bts163

S45.	Villicaña S, Castillo-Fernandez J, Hannon E, Christiansen C, Tsai PC, Maddock J, et al. Genetic impacts on DNA methylation help elucidate regulatory genomic processes. *Genome Biol*. 2023;24(1):176. doi:10.1186/s13059-023-03011-x

S46.	Bonder MJ, Luijk R, Zhernakova DV, Moed M, Deelen P, Vermaat M, et al. Disease variants alter transcription factor levels and methylation of their binding sites. *Nat Genet*. 2017;49(1):131-138. doi:10.1038/ng.3721

S47.	Ruiz-Arenas C, Hernandez-Ferrer C, Vives-Usano M, Marí S, Quintela I, Mason D, et al. Identification of autosomal cis expression quantitative trait methylation (cis eQTMs) in children’s blood. *eLife*. 2022;11:e65310. doi:10.7554/eLife.65310

S48.	Perez-Garcia J, Cardenas A, Lorenzo-Diaz F, Pino-Yanes M. Precision medicine for asthma treatment: Unlocking the potential of the epigenome and microbiome. *J Allergy Clin Immunol*. 2025;155(2):298-315. doi:10.1016/j.jaci.2024.06.010

S49.	Xu Z, Niu L, Kresovich JK, Taylor JA. methscore: a comprehensive R function for DNA methylation-based health predictors. Alkan C, ed. *Bioinformatics*. 2024;40(5):btae302. doi:10.1093/bioinformatics/btae302

### SUPPLEMENTARY FIGURES

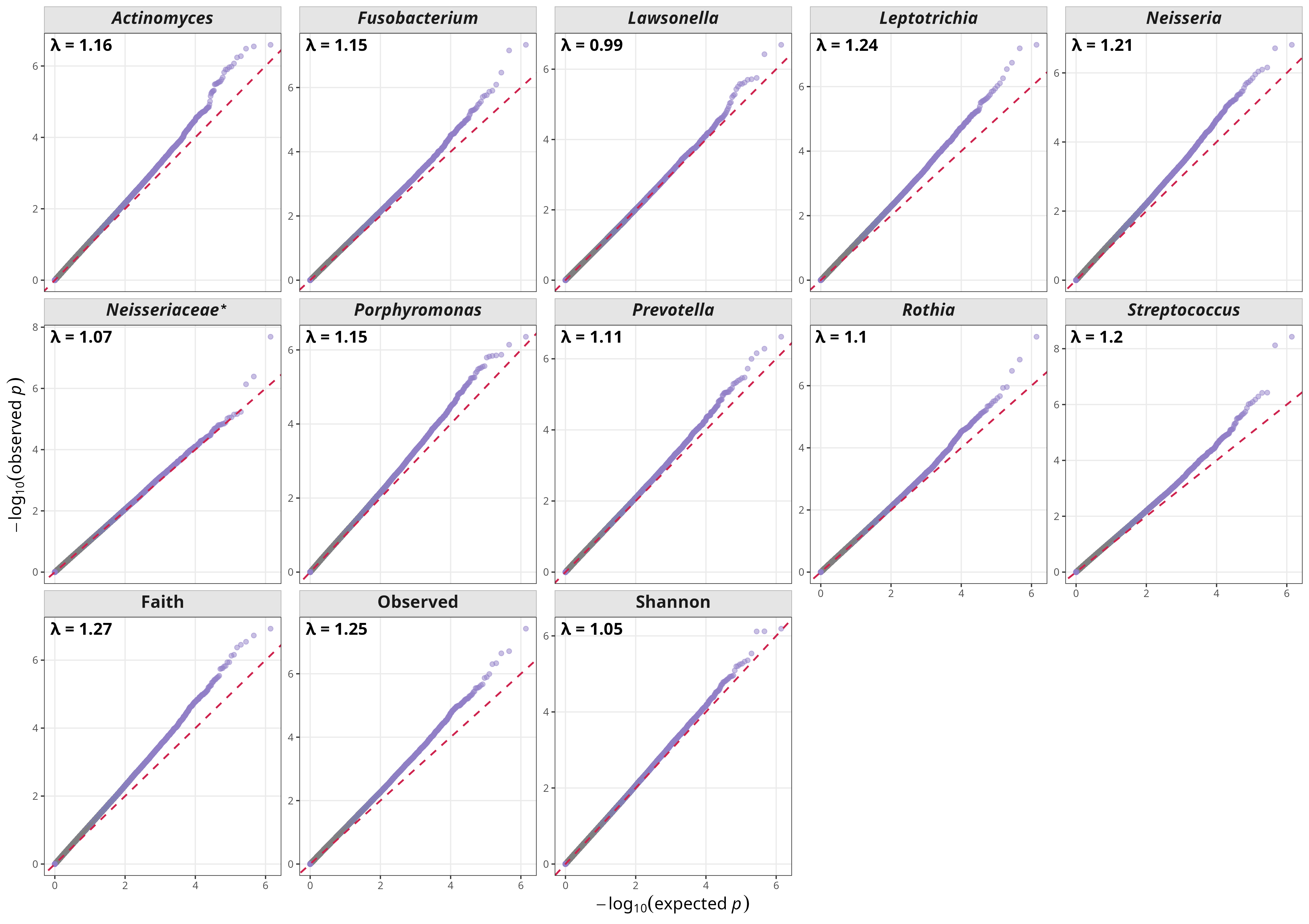

**Figure S1**. Quantile-Quantile (Q-Q) plots for the nasal mbEWAS in the discovery cohort, shown separately for each bacterial genus and α-diversity index tested. The x-axis shows expected -log_10_(p-values) under the null hypothesis of no association, and the y-axis shows observed -log_10_(p-values). The red diagonal indicates the expected distribution under the null hypothesis. *The lowest taxonomy classification available was at the family level. Observed: Observed richness; Shannon: Shannon diversity index; Faith: Faith’s phylogenetic diversity index.

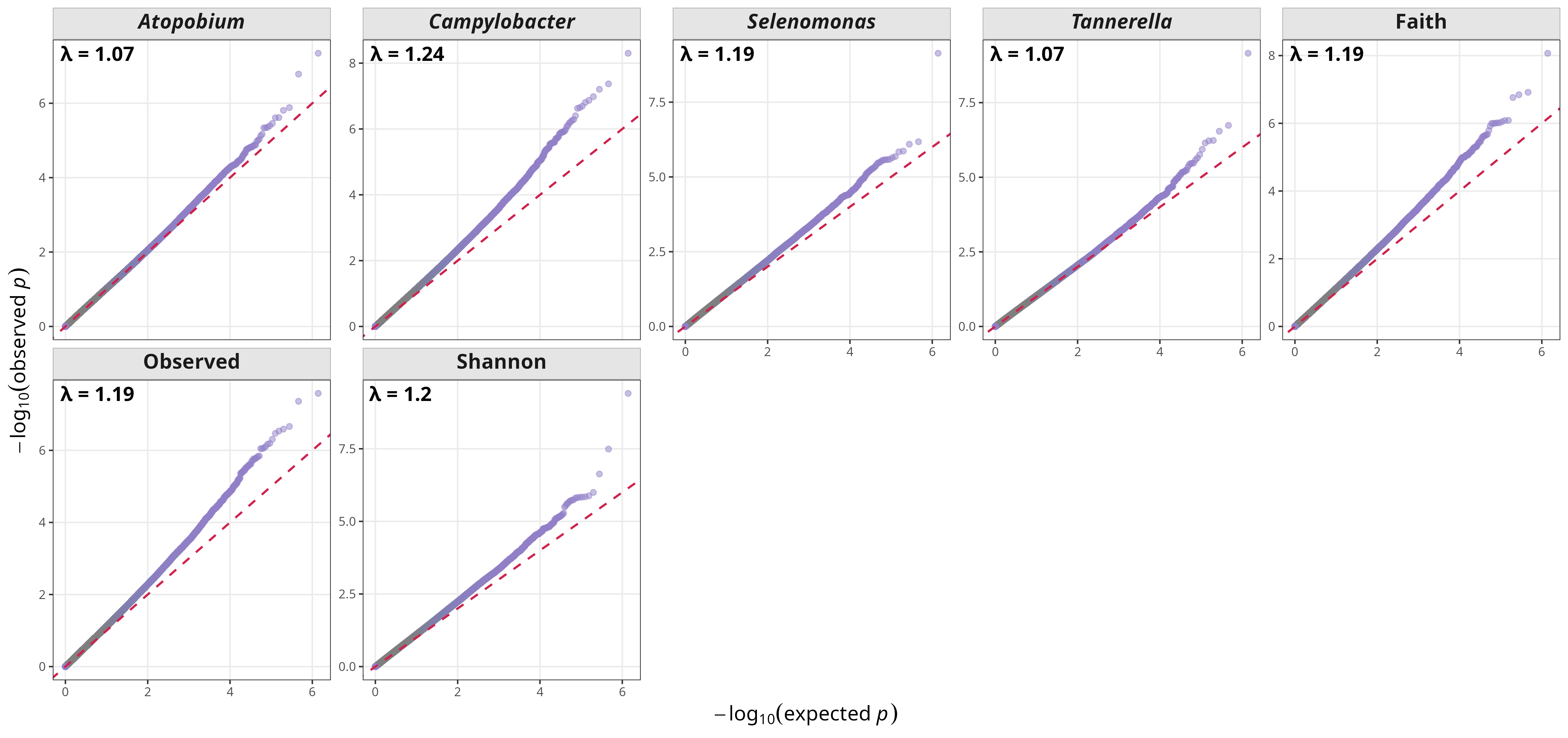

**Figure S2**. Quantile-Quantile (Q-Q) plots for the pharyngeal mbEWAS in the discovery cohort, shown separately for each bacterial genus and α-diversity index tested. The x-axis shows expected -log_10_(p-values) under the null hypothesis of no association, and the y-axis shows observed -log_10_(p-values). The red diagonal indicates the expected distribution under the null hypothesis. Observed: Observed richness; Shannon: Shannon diversity index; Faith: Faith’s phylogenetic diversity index.

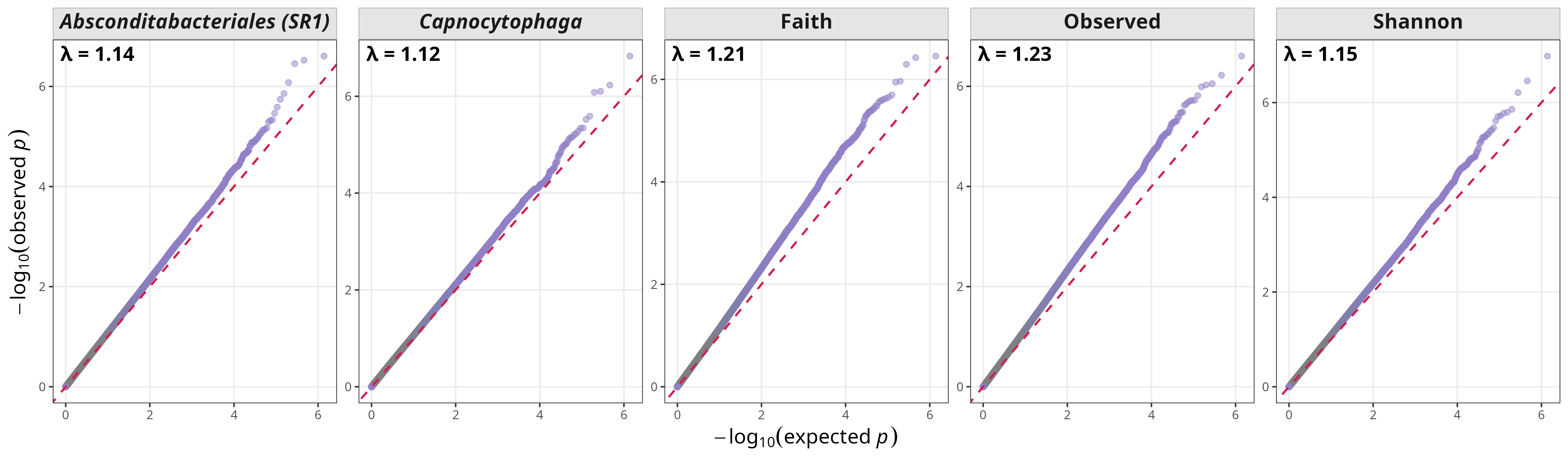

**Figure S3**. Quantile-Quantile (Q-Q) plots for the salivary mbEWAS in the discovery cohort, shown separately for each bacterial genus and α-diversity index tested. The x-axis shows expected -log_10_(p-values) under the null hypothesis of no association, and the y-axis shows observed -log_10_(p-values). The red diagonal indicates the expected distribution under the null hypothesis. Observed: Observed richness; Shannon: Shannon diversity index; Faith: Faith’s phylogenetic diversity index.

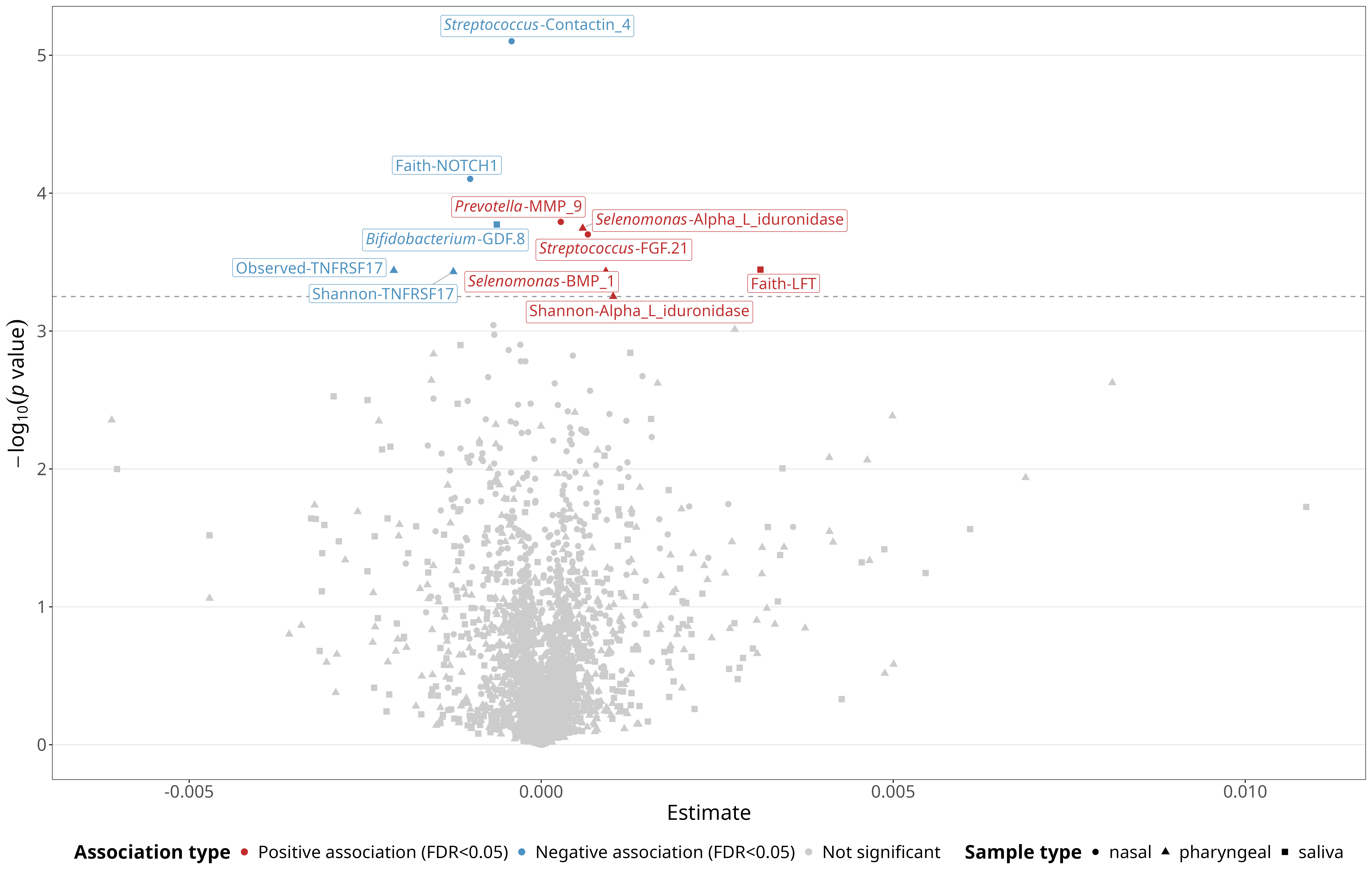

**Figure S4**. Volcano plot of DNAm-predicted plasma proteins associated with the exacerbation-related upper-airway microbiome. Association −log_10_(p) from linear regression models is represented on the y-axis and effects on the x-axis. Only significant associations (FDR<0.05) are colored in blue (negative association) and red (positive association). The dashed line indicates the significance threshold. Shapes indicate the sample type: nasal (circles), pharyngeal (triangles), and saliva (squares). Observed: Observed richness; Shannon: Shannon diversity index; Faith: Faith’s phylogenetic diversity index.
